# Epidemiological dynamics of the incidence of COVID-19 in children and the relationship with the opening of schools in Catalonia (Spain)

**DOI:** 10.1101/2021.02.15.21251781

**Authors:** Aida Perramon, Antoni Soriano-Arandes, David Pino, Uxue Lazcano, Cristina Andrés, Martí Català, Anna Gatell, Mireia Carulla, Dolors Canadell, Gemma Ricós, M<u>^a^</u> Teresa Riera-Bosch, Silvia Burgaya, Olga Salvadó, Javier Cantero, Mònica Vilà, Miriam Poblet, Almudena Sánchez, Anna M<u>^a^</u> Ristol, Pepe Serrano, Andrés Antón, Clara Prats, Pere Soler-Palacin, on behalf of COPEDI-CAT research group

**Affiliations:** Universitat Pompeu Fabra, Barcelona, Catalonia, Spain; These authors contributed equally to this article and share first authorship; Paediatric Infectious Diseases and Immunodeficiencies Unit, Hospital Universitari Vall d’Hebron, Barcelona, Catalonia, Spain; Department of Physics, Universitat Politècnica de Catalunya (UPC·BarcelonaTech), Barcelona, Catalonia, Spain; Agència de Qualitat i Avaluació Sanitàries de Catalunya (AQuAS), Barcelona, Catalonia, Spain; Respiratory Viruses Unit, Department of Microbiology, Hospital Universitari Vall d’Hebron, Barcelona, Catalonia, Spain; Comparative Medicine and Bioimage Centre of Catalonia (CMCiB), Fundació Institut d’Investigació en Ciències de la Salut Germans Trias i Pujol (IGTP), Badalona, Catalonia, Spain; Equip Pediatria Territorial Alt Penedès-Garraf, Barcelona, Catalonia, Spain; ABS Pla d’Urgell (Mollerussa), Lleida, Catalonia, Spain; CAP Barberà del Vallés, Barcelona, Catalonia, Spain; CAP Drassanes, Barcelona, Catalonia, Spain; EAP Vic Nord, Barcelona, Catalonia, Spain; EAP Manlleu, Barcelona, Catalonia, Spain; CAP Llibertat Reus, Tarragona, Catalonia, Spain; Corporació del Maresme i la Selva, Barcelona, Catalonia, Spain; EAP Horta, Barcelona, Catalonia, Spain; Equip Territorial Pediàtric Sabadell Nord, Barcelona, Catalonia, Spain; CAP Les Hortes, Barcelona, Catalonia, Spain; CAP Can Serra Hospitalet de Llobregat, Barcelona, Catalonia, Spain

**Keywords:** COVID-19, SARS-CoV-2, child, epidemiology, school

## Abstract

Here we analyse the epidemiological trend of the incidence of COVID-19 in children in Catalonia (Spain) during the first 20 weeks of the 2020-2021 school year. This study demonstrates that while schools were open the incidence rate among children remained significantly lower than in general population, despite a greater diagnostic effort in children. These results suggest that schools have not played a significant role in the SARS-CoV-2 dissemination in Catalonia.

## ARTICLE

During the COVID-19 pandemic, the role of schools in viral transmission among children [1] and the impact of the school closures on children [2] have been at the centre of public debate. Some meta-analyses and review studies analysing household transmission have shown that children and adolescents are less susceptible than adults to severe acute respiratory syndrome coronavirus 2 (SARS-CoV-2) infection [3, 4], and that the dynamics of the epidemic are mainly driven by the 20-49 age-group [5]. However, differences may exist between kindergarten, primary, and secondary age groups [6].

We describe and analyse here the epidemiological trend of childhood (<18 years of age) COVID-19 incidence in Catalonia (Spain) during the first 20 weeks of the 2020-2021 school year (14^th^ September to 31^st^ January), including the Christmas holidays period (22^nd^ December to 10^th^ January). We focused our research on age-group incidence, diagnostic effort, and positivity percentage; these indicators were compared with those for the general population. A confirmed COVID-19 case was defined as any individual testing SARS-CoV-2 positive by molecular assays (PCR or TMA-based) or rapid antigen testing (RAT) (PANBIO COVID-19 by ABBOTT©) in a respiratory specimen. RAT was only available from 23^rd^ October 2020 in primary care settings. All the contacts of the confirmed cases were tested only through PCR.

Catalonia is an autonomous region in north-eastern Spain with 7,653,845 inhabitants (1,384,382 < 18 years). We gathered data of the total tested and confirmed SARS-CoV-2 cases in Catalonia delivered by the Catalan Agency for Quality and Health Assessment (AQuAS) that had downloaded the original data from the Catalan Epidemiological Surveillance Network’s clinical microbiological laboratories [7]. Schools were re-opened in Catalonia on 14^th^ September 2020, after a long closure period, from 13^th^ March 2020. This reopening occurred in a context in which the average 14-day cumulative incidence was around 180-190 cases per 10^5^ inhabitants, and the most prevalent SARS-CoV-2 Pangolin lineage was B.1.177 with other minor lineages. The planning for the schools included a set of non-pharmaceutical interventions (NPI) to minimize the risk of transmission and increase the diagnostic level. This may be summarized as follows: 1) mask wearing was mandatory in all the educational centres for children from 6 years old on, 2) natural ventilation, hand hygiene and infographic about COVID-19 in the classrooms and common spaces was recommended, 3) infants and adolescents were clustered in bubble-groups (mainly restricted to classmates) in all educational centres with no interactions with other groups in the centre, and 4) whenever a positive case was detected, the whole group was screened and, whatever the result of the SARS-CoV-2 tests, quarantining was established for the whole group for 10 days. In addition, several mass screening campaigns were carried out in educational centres located in high incidence areas. During the analysed period, the regional government introduced various NPIs to reduce the global incidence of COVID-19 (**Supplementary Table S1**).

During the study period, 942,881 SARS-CoV-2 tests were performed in the population < 18 years, and 48,914 of them were positive (5.2%). **Figure 1** shows the daily incidence per 10^5^ inhabitants in each age-group according to the different educational age groups (0-2, 3-5, 6-11, 12-17 years), and comparison with the overall incidence in Catalonia. The incidence for children <18 years was always lower than the incidence for the general population during the study-period except during late January 2021. Among children, similarly to other studies [8, 9], higher incidence was associated with the age of the group, but the evolution of incidence for all age-groups followed a similar pattern. Additionally, the incidence among children <12 years remained at a lower level when compared to the general population, while the 12-17 age-group remained at a similar level or higher.

Nevertheless, incidence could have been affected by changes in the diagnostic effort. To avoid this bias, we show weekly 7-day cumulative incidence and weekly COVID-19 laboratory tests per 10^5^ people for each age-group and for the general population (**Figure 2**). Up to the end of November, the diagnostic effort in children (except for <3 years) was greater than for the general population mainly due to the active surveillance screening when a positive case occurred in a school bubble-group. This diagnostic effort was especially large for the age-group 12-17 during the first 11 weeks of the school year and then after the Christmas holidays. In contrast, during Christmas holidays, while schools were closed, this active screening was not performed and, consequently, the number of tests performed for all age-groups was much closer to the standard for the general population, limited to high-risk contacts.

A relevant variable when trying to combine the incidence with the diagnostic effort is the percentage of positive samples among tests performed, as the so-called positivity rate. **Figure 3** shows the evolution of the weekly positivity rate by age-group and among the general population. During the first 11 weeks of the school year, positivity among children remained clearly below the Catalan average, with percentages around 5% or even lower in younger children. This fact indicates, at least in part, that the higher incidence seen in **Figure 1** for the 12-17 age-group could be explained by a greater effort at diagnosis due to mass testing. In contrast, after the long weekend at the beginning of December and then again during the Christmas holidays, the positivity percentage for all child age-groups increased substantially, because of a low diagnosis effort as shown in **Figure 2**. At the end of December the first SARS-CoV-2 cases related to the B.1.1.7 lineage were detected in Catalonia, co-circulating with the more prevalent B.1.177 viruses; prevalence of B.1.1.7 increased to 17% by week 05/2021 (unpublished data).

We performed an ANOVA test to explore differences in tests, cases, and positivity between 0-11 years, 12-17 years, and adults, in comparison with the data for the general population. To detect all real cases linked to the period when the schools were closed, the analysis of cases and positivity defined this period as from one week after the beginning of the Christmas holidays (28^th^ December) to one week after returning to school (17^th^ January). Daily tests and cases among children and adolescents compared with the general population decreased significantly when the schools were closed (p < 0.001) **(Figure 4)**. Moreover, the number of cases < 18 years divided by the number of cases in the general population was significantly lower than for adults during the whole study period (p < 0.001). Finally, positivity increased when the schools were closed mainly due to a decrease in the diagnostic effort (p < 0.001).

## Conclusions

This study shows that, while schools were re-opened and appropriate NPIs were implemented, global COVID-19 incidence among children remained lower than the incidence for the general population despite the greater diagnostic effort in this age-group. However, a distinction should be made between children attending kindergarten or primary schools (0-11 years), and adolescents in secondary/high schools (12-17 years) [10]. Incidence positivity rates per age-group suggest that the former have not played a significant role in SARS-CoV-2 dissemination in Catalonia probably due to their lower susceptibility to the virus and lesser capacity to transmit it [4, 9]. Since the novel SARS-CoV-2 variants belonging to the B.1.1.7 and B.1.351 lineages might be characterized by higher transmissibility and may also become the most prevalent viruses in the coming weeks, schools are a perfect scenario for monitoring the pandemic among children and key to initiating contact-tracing studies among students’ families.

**Figure 1.**
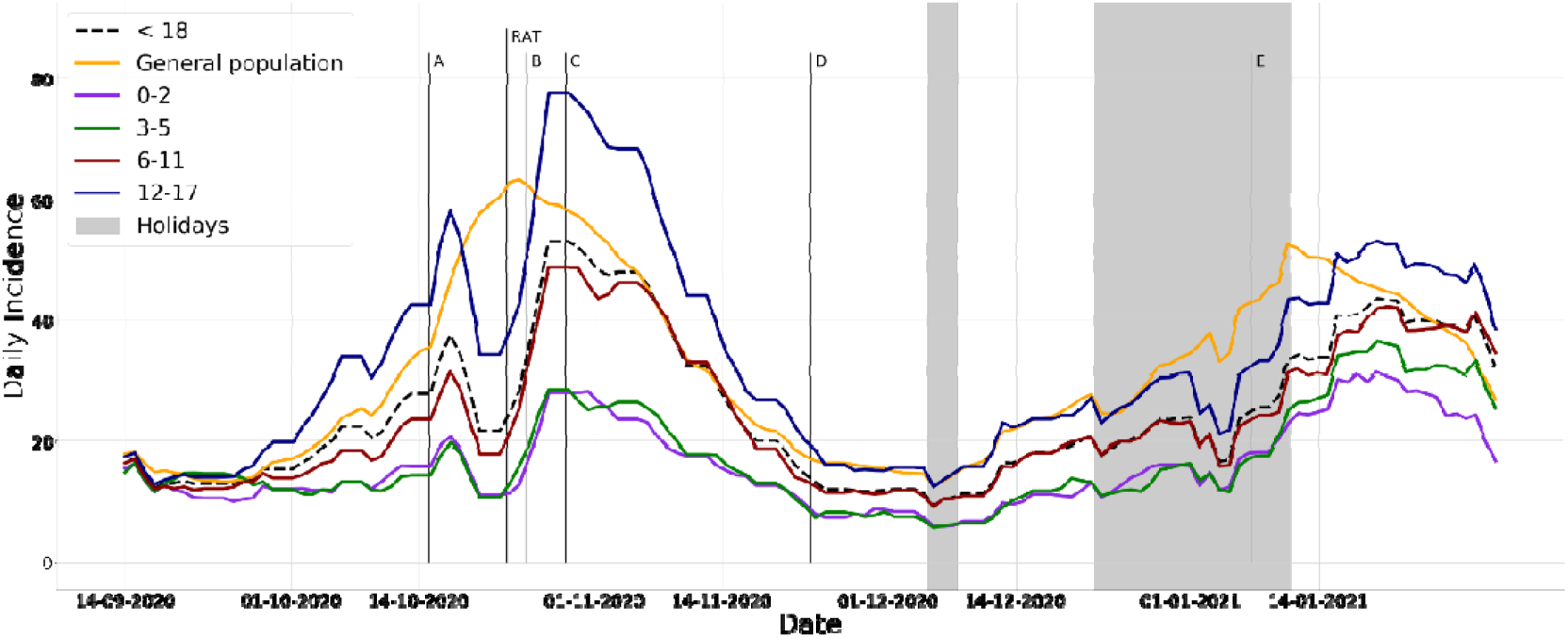
Daily positive cases per 10^5^ population for each age cohort, and for the general population. A 7-day running average was applied to rough data. Letters A-E indicate when the different NPIs were implemented. RAT indicates when rapid antigen tests were introduced.

**Figure 2.**
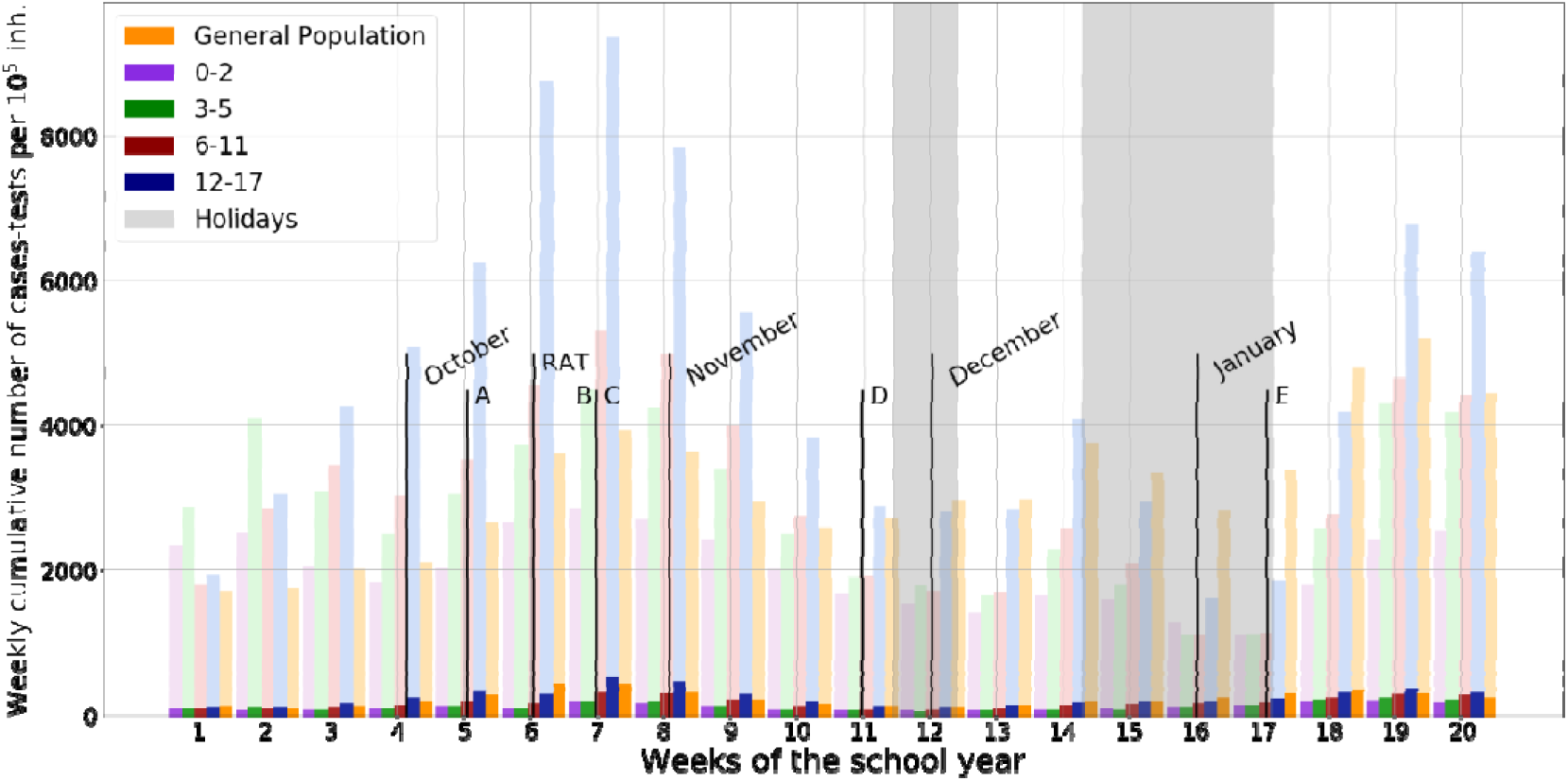
Weekly cumulative number of tests (light) and cases (dark) per 10^5^ population for each age cohort and for the general population. Letters A-E indicate when the different NPIs were implemented. RAT indicates when rapid antigen tests were introduced.

**Figure 3.**
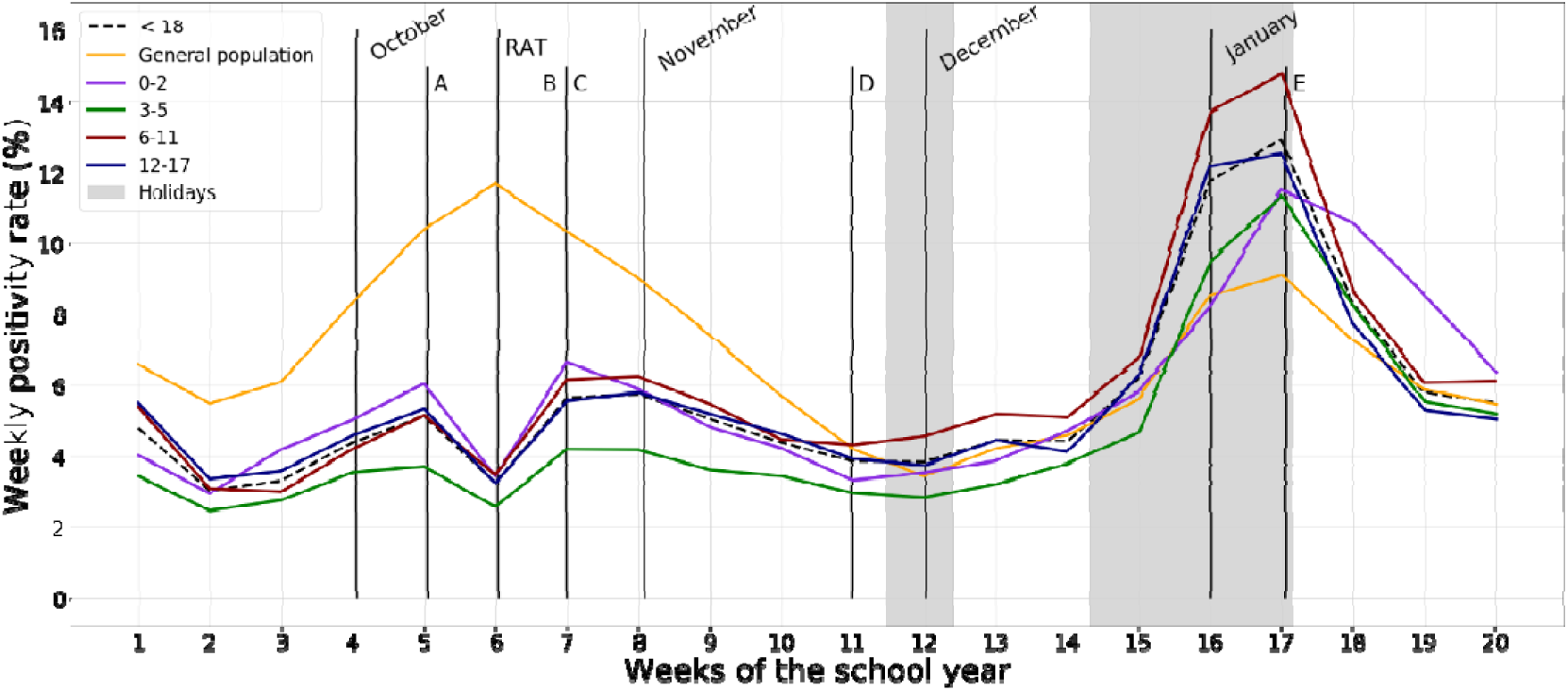
Weekly positivity (percentage of positive cases among tested population) for each age cohort and for the general population). Letters A-E indicate when the different NPIs were implemented. RAT indicates when rapid antigen tests were introduced.

**Figure 4.**
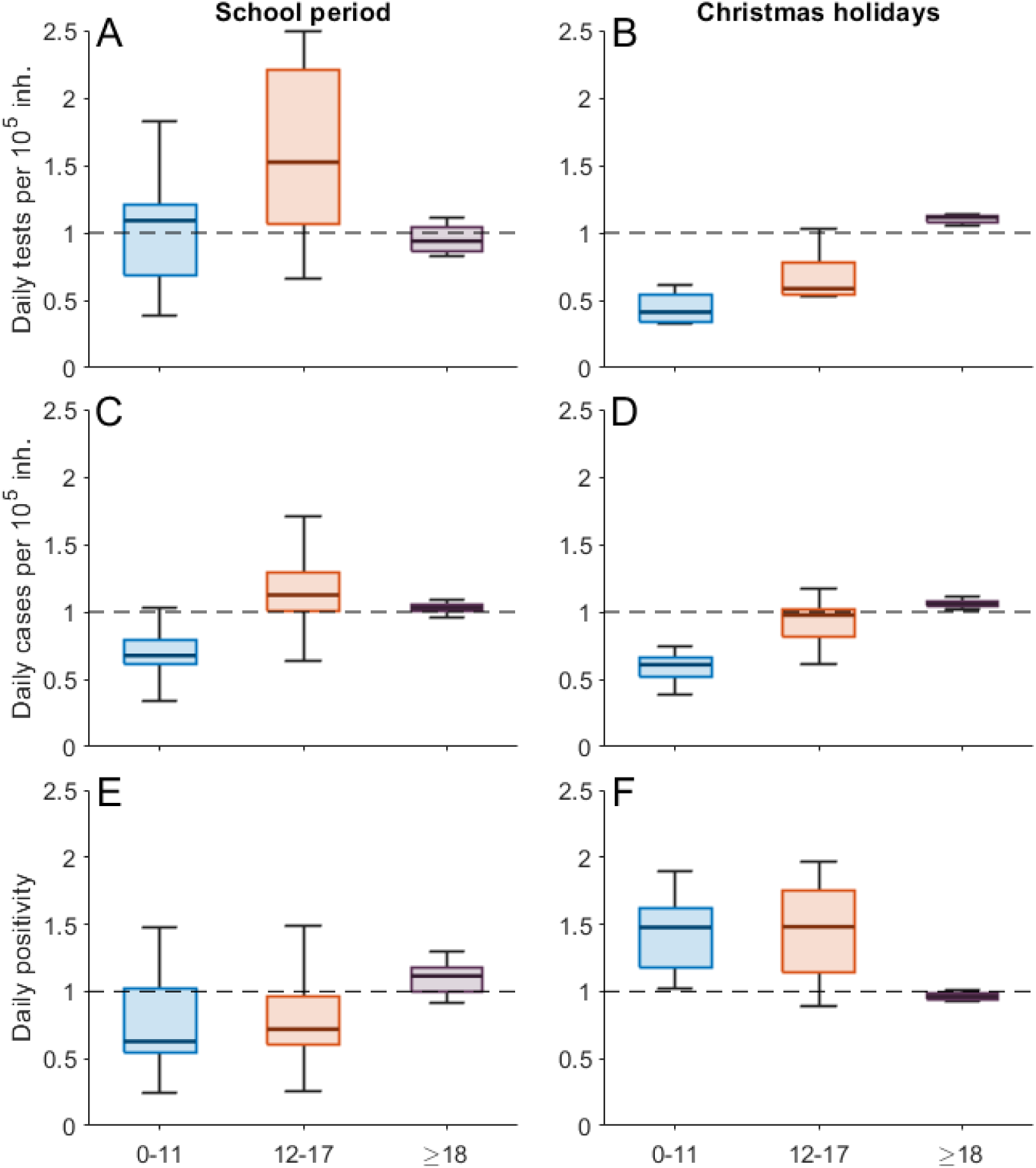
Box plots for the daily number of tests performed per 10^5^ inhabitants (top), daily incidence (middle), and daily positivity (bottom) during the school period (left) and holidays (21-12-2020/10-01-2021 for tests and 28-12-2020/17-01-2021 for cases and positivity, right). For each age group, the variables are presented relative to the general population (i.e., value in the age group divided by the value in the general population).

## Data Availability

The data referred to in the manuscript is available from the Catalan Agency for Health Quality and Evaluation (AQuAS)

## Ackowledgments

This study was partially supported by the Direcció General de Recerca i Innovació en Salut (DGRIS), Catalan Health Ministry, Generalitat de Catalunya through Vall d’Hebron Research Institute (VHIR).

