## Supplementary table S1 for "Epidemiological dynamics of the incidence of COVID-19 in children and the relationship with the opening of schools in Catalonia (Spain)"

**Supplementary Table S1**. Non-pharmaceutical interventions implemented by the Autonomous Government during the analysed period.

|  | Description | Date of application | Details |
| --- | --- | --- | --- |
| A | 1^st^ set of NPI | 15-10-2020 | - Social activities: - Meetings of < 6 people - Between “cohabitation bubbles” and “extended bubbles” - Limited non-essential travels - Cultural and leisure activities: - Closed: bingos, casinos, arcades, children's indoor play areas, local festivals - 50% capacity in cultural activities and preassigned seats - Closing time at 23:00 h - Children’s playgrounds open until 20:00 h - Shops and shopping centres: - 30% capacity in retail businesses - 1,5 m of minimum distance between customers - Closed: Beauty centres, except from hairdressers - 30% capacity in non-sedentary markets - Religious acts and civil ceremonies: - 50% capacity - Sports: - 50% capacity in gyms - Closed: unsupervised sports venues - Postponed: non-professional competitions - Not postponed: professional competitions - Hotels and restaurants: - Closed: only home delivery or pick-up at the establishment by appointment - 50% capacity in common areas in hotels - Universities and schools: - Opened: schools, institutes, extracurricular activities and sports - Universities: online theoretical teaching - Companies: - Promotion of telework - Air renovation before and after working hours - Suspended: congresses, conventions and trade fairs |
| B | Introduction of RAT | 23-10-2020 | - Introduction of RAT in addition to PCR tests |
| C | 2^nd^ set of NPI | 25-10-2020 | - Night-time confinement, from 22:00 h to 6:00 h - Closed to the public at 21:00 h: services, retail businesses, restaurants, sportive and recreational activities and public spaces - Closed to the public at 22:00 h: cultural activities. Attendees are allowed to return home between 22:00 h and 23:00 h - Exceptions: - Emergency healthcare - Acquisition of pharmaceutical products as a matter of urgency - Travel to and from work, with the corresponding company certificate - Travel of professionals, and accredited volunteer staff, to provide essential, health and social services - Care for the elderly, minors, dependents either disabled or especially vulnerable, for undelayable reasons - Urgent action in the courts - Return to the place of usual residence - Pet urgent care from 4:00 h to 6:00 h, individual travel - Other causes of justified need |
| D | 3^rd^ set of NPI | 29-10-2020 | - Forbidden to enter or leave Catalonia - During the weekends, forbidden to enter or leave municipality from 6:00 h on Friday until 6:00 h on Monday. Exceptions: - Visits to the region’s cemeteries on October 31^st^ and All Saints’ Day - Health services, work trips or causes of justified need - Individual sports activities between neighbouring municipalities - Closure of bars and restaurants extended. Food delivery at home until 23:00 h - Only loan service in libraries - Closed: theatres, cinemas, concert halls, sports centres and gyms. - Shopping centres, except for shops that are food and essential services - Shops of more than 800 m^2^ if the area is not reduced to 800 m^2^ and limit the capacity to 30% - Extracurricular activities, except those done at school keeping the “group bubble” |
| E | Relaxation of previous NPI | 23-11-2020 | - Social activities: - Meetings of < 6 people, within the “usual bubble” - Cultural and leisure activities: - Closed: bingos, casinos, arcades, children's indoor play areas, local festivals - 50% capacity in museums, libraries and exposition halls - 50% capacity and 500 people limit in cinemas, theatres, auditoriums and concert halls - Children’s playgrounds open until 20:00 h - Shops and shopping centres: - Closed: shopping centres, except from first need shops with public access - Opened: establishments offering close physical contact services, including hairdressers and beauty centres, with prior appointment and preventive measures - 30% capacity in shops and non-sedentary markets - Religious acts: - 30% capacity - Sports: - 50% capacity in outdoor sports facilities and equipment - 30% capacity in indoor facilities, with prior appointments and no dressing rooms, except for swimming activities - Restaurants: - Home delivery until 23:00 h or pick-up at the establishment until 22:00 h - Terraces open from 6:00 h to 21:30 h - 30% capacity in indoor spaces from 6:00 h to 21:30 h - 4 people per table and 2 m of separation between tables - Extracurricular activities: - Opened: non-competitive sport activities in school, leisure educational activities outdoors - Maximum of 6 people in indoor leisure educational activities and on-site extracurricular activities - Education: - Opened: schools and institutes - Reduction of on-site activities: baccalaureate, training courses and regulated general education courses - Universities: online theoretical teaching - Companies: - Promotion of telework - Suspended: congresses, conventions and fairs - Lockdown: - Forbidden to enter or leave Catalunya without justified motivation - Forbidden to enter or leave the municipality from Friday at 6:00 h to Monday at 6:00 h - Night-time confinement, from 22:00 h to 6:00 h, except from justified needs |
| F | January NPI | 07-01-2021 | - Social activities: - Meetings of < 6 people, from two the “cohabitation bubbles” - Indoor meetings limited to visits to dependent or vulnerable people within the “cohabitation bubble” - Cultural and leisure activities: - Closed: bingos, casinos, arcades, children's indoor play areas, local festivals - 50% capacity in cinemas, theatres, auditoriums, concert halls, museums, libraries and exposition halls. 500 people limit indoors, 1000 people limit outdoors - 50 % capacity in children’s playgrounds, opened until 20:00 h - Shops and shopping centres: - Closed: shopping centres and shops of more than 400 m^2^, except first need establishments - 30% capacity in retail businesses, shopping centres of less of 400 m^2^ and non-sedentary markets. Opened from 6:00 h to 21:00 h - Religious acts: - 30% capacity, maximum of 500 people indoors, 1000 people outdoors - Sports: - 50% capacity in outdoor sports facilities and equipment, < 6 people - Closed: indoor facilities and activities, competitions - Restaurants: - Home delivery until 23:00 h or pick-up at the establishment until 22:00 h - Terraces open from 7:30 h to 09:30 h and from 13:00 h to 15:30 h - 30% capacity in indoor spaces from 7:30 h to 09:30 h and from 13:00 h to 15:30 h - 4 people per table and 2 m of separation between tables - Extracurricular and leisure activities: - Opened: activities in school or organised by educational centres, within the “educational group bubble” - Closed: all others - Education: - Opened: schools and institutes - Reduction of on-site activities: baccalaureate, training courses and regulated general education courses - Universities: online theoretical teaching - Companies: - Promotion of telework - Suspended: congresses, conventions and fairs - Lockdown: - Forbidden to enter or leave Catalunya without justified motivation - Forbidden to enter or leave the municipality without justified reason or to work - Night-time confinement, from 22:00 h to 6:00 h, except from justified needs |
